# Population Age-Ineligible for COVID-19 Vaccine in the United States: Implications for State, County, and Race/Ethnicity Vaccination Targets

**DOI:** 10.1101/2021.02.11.21251562

**Authors:** Elizabeth B. Pathak, Janelle Menard, Rebecca Garcia

## Abstract

**Background:** We examined the geographic and racial/ethnic distribution of the SARS-CoV-2 vaccine age-ineligible population (0-15 years old) in the U.S., and calculated the proportion of the age-eligible population that will need to be vaccinated in a given geo-demographic group in order to achieve either 60% or 75% vaccine coverage for that population as a whole.

**Methods:** US Census Bureau population estimates for 2019 were used to calculate the percent vaccine ineligible and related measures for counties, states, and the nation as a whole. Vaccination targets for the 30 largest counties by population were calculated. Study measures were calculated for racial/ethnic populations at the national (n=7) and state (n=6) levels.

**Results:** Percent of population ineligible for vaccine varied widely both geographically and by race/ethnicity. State values ranged from 15.8% in Vermont to 25.7% in Utah, while percent ineligible of the major racial/ethnic groups was 16.4% of non-Hispanic whites, 21.6% of non-Hispanic Blacks, and 27.5% of Hispanics. Achievement of total population vaccine coverage of at least 75% will require vaccinating more than 90% of the population aged 16 years and older in 29 out of 30 of the largest counties in the U.S.

**Conclusions:** The vaccine-ineligibility of most children for the next 1-2 years, coupled with reported pervasive vaccine hesitancy among adults, especially women and most minorities, means that achievement of adequate levels of vaccine coverage will be very difficult for many vulnerable geographic areas and for several racial/ethnic minority groups, particularly Hispanics, Blacks, and American Indians.

## Background

The demographic structure of populations is an important constraint on COVID-19 vaccination efforts. Unlike many of the aging countries of western Europe, the United States has a large child population. In 2019, there were 64.4 million children aged 0 to 15 years in the United States, comprising 19.7% of the total population.^1^ Children less than 16 years old are ineligible to receive either of the two vaccines authorized for emergency use in the United States,^2,3^ and there are no vaccines for children that have completed any clinical trials.^4,5^ A third vaccine, which may receive FDA emergency use authorization in February 2021, has also been trialed only in adults.^6^ Late 2021 is the earliest timeframe that a vaccine for children will likely be approved and ready for mass distribution.^7^ Even then, the first child vaccines will be approved only for ages 12 to 15 years old.^5,7^

There is no known inborn immunity to SARS-CoV-2 for children of any age. More than 3.2 million (as of 21-Jan-21) infants, children, and teens have tested positive for the virus in the US.^8^ Many thousands have been hospitalized and several thousand have been admitted to intensive care critically ill with COVID-19.^8^ Despite popular disbelief in some quarters, there is robust scientific evidence from a wide range of settings that the novel coronavirus can be transmitted both from children to other children and to adults.^9-15^

Consequently, aspirational plans to achieve population herd immunity to SARS-CoV-2 through widespread vaccination must consider the vaccine-ineligible child population. The purpose of this study was to examine the geographic and racial/ethnic distribution of the SARS-CoV-2 vaccine age-ineligible population in the U.S., and to calculate the proportion of the age-eligible population that would need to be vaccinated in a given geographic area or racial/ethnic group in order to achieve either 60% or 75% vaccine coverage for the population as a whole.

## Methods

Intercensal population estimates for 2019, the latest year available, were obtained from CDC WONDER.^1^ For national and state estimates, single-year of age population counts were used to derive the population 0-15 years old. For counties, single-year of age estimates were unavailable. Therefore, for every county we obtained population estimates for the following 5-year age groups: <1 year, 1-4 years, 5-9 years, 10-14 years, and all ages combined. County populations 0-15 years old were estimated using the following formula:

(2 x population <1 year) + (population 1-4 years) + (population 5-9 years) + (population 10-14 years) The following variables were defined for our analyses:

> <u>Proportion of the population ineligible for vaccine</u> = population 0-15 years old / total population.
>
> <u>Proportion eligible for vaccine</u> = 1 – proportion ineligible.
>
> <u>Proportion of the eligible population who would need to be vaccinated to achieve 60% vaccine coverage for the total population</u> = .60 / proportion eligible.
>
> <u>Proportion of the eligible population who would need to be vaccinated to achieve 75% vaccine coverage for the total population</u> = .75 / proportion eligible.

We calculated these measures for the nation, states, and counties, and for racial/ethnic population groups at the national and state level. Population estimates for the following racial/ethnic groups were analyzed: non-Hispanic whites, Hispanics of any race, non-Hispanic Blacks, non-Hispanic Asians, non-Hispanic American Indians and Alaska Natives, non-Hispanic Native Hawaiians and Alaska Natives, non-Hispanic persons reporting more than one race. We also report findings for the Hispanic American Indian and Alaska Native population, because 47% of American Indian/Alaska Native children in 2019 were also Hispanic.

## Results

Nationally, vaccine-ineligible children aged 0 to 15 years old are 19.7% of the total population. For states, the percent of population ineligible for vaccine ranges from 15.8% in Vermont to 25.7% in Utah (Figure 1). Among the 3,142 counties, percent ineligible ranges from 0% (1 county with child population counts suppressed due to very low numbers) to 38.3%, with a median value of 19.2%, and 1^st^ to 99^th^ percentile range from 11.9% to 29.0%.

**Figure 1.**
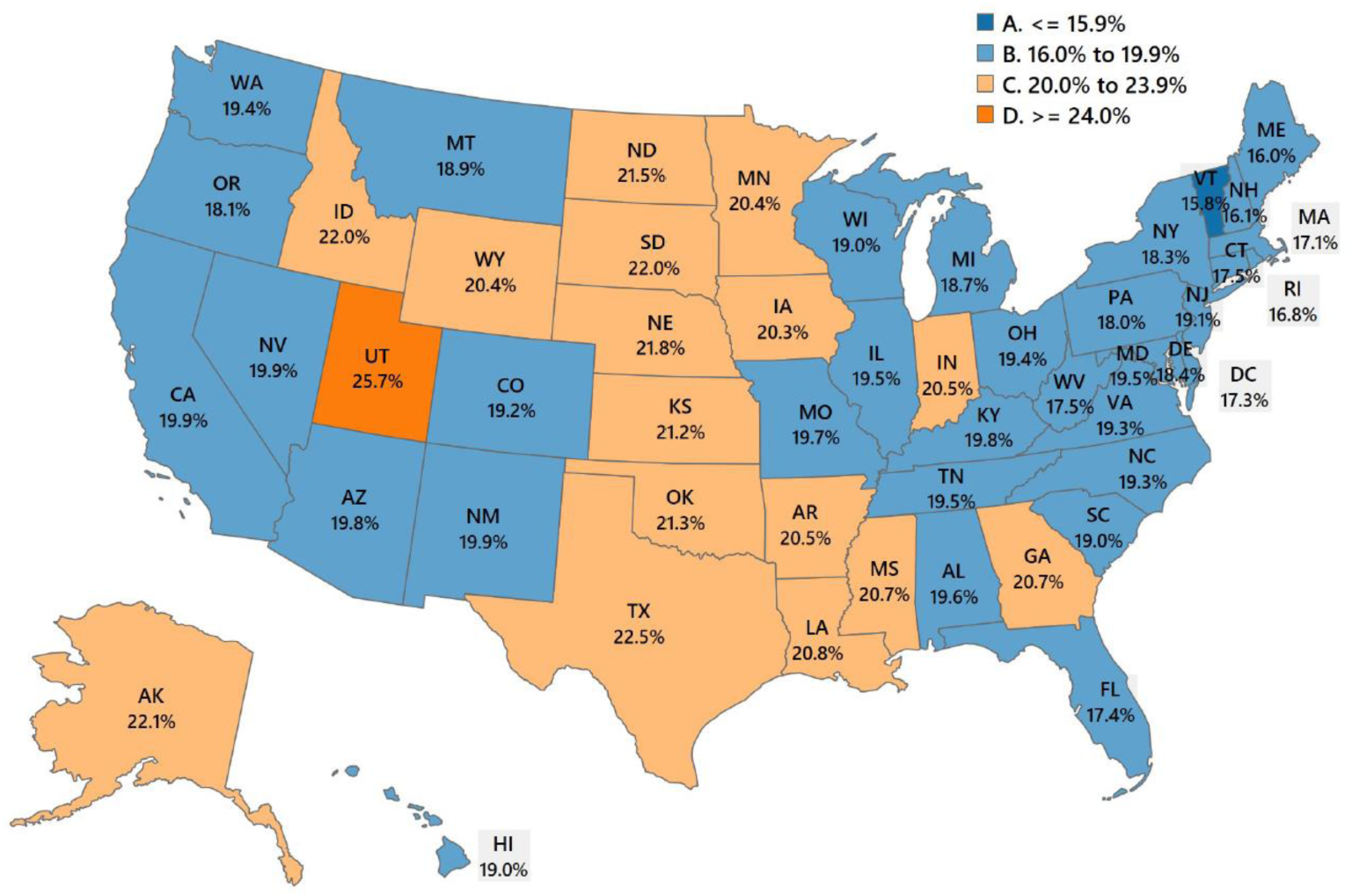
Percent of Total Population Who Are Age-Ineligible for Covid-19 Vaccine in U.S C States. *(percent of population aged 0 to 15 years old)*

Local variation in percent ineligible is shown in Figure 2. Appalachia, New England, and Florida stand out as regions with proportionately smaller child populations. In contrast, the Rio Grande Valley and West Texas, California’s Central Valley, the Mississippi Delta, Utah, arctic Alaska, and many tribal areas of the West stand out as local regions with age-ineligible populations greater than 24% of the total population.

**Figure 2.**
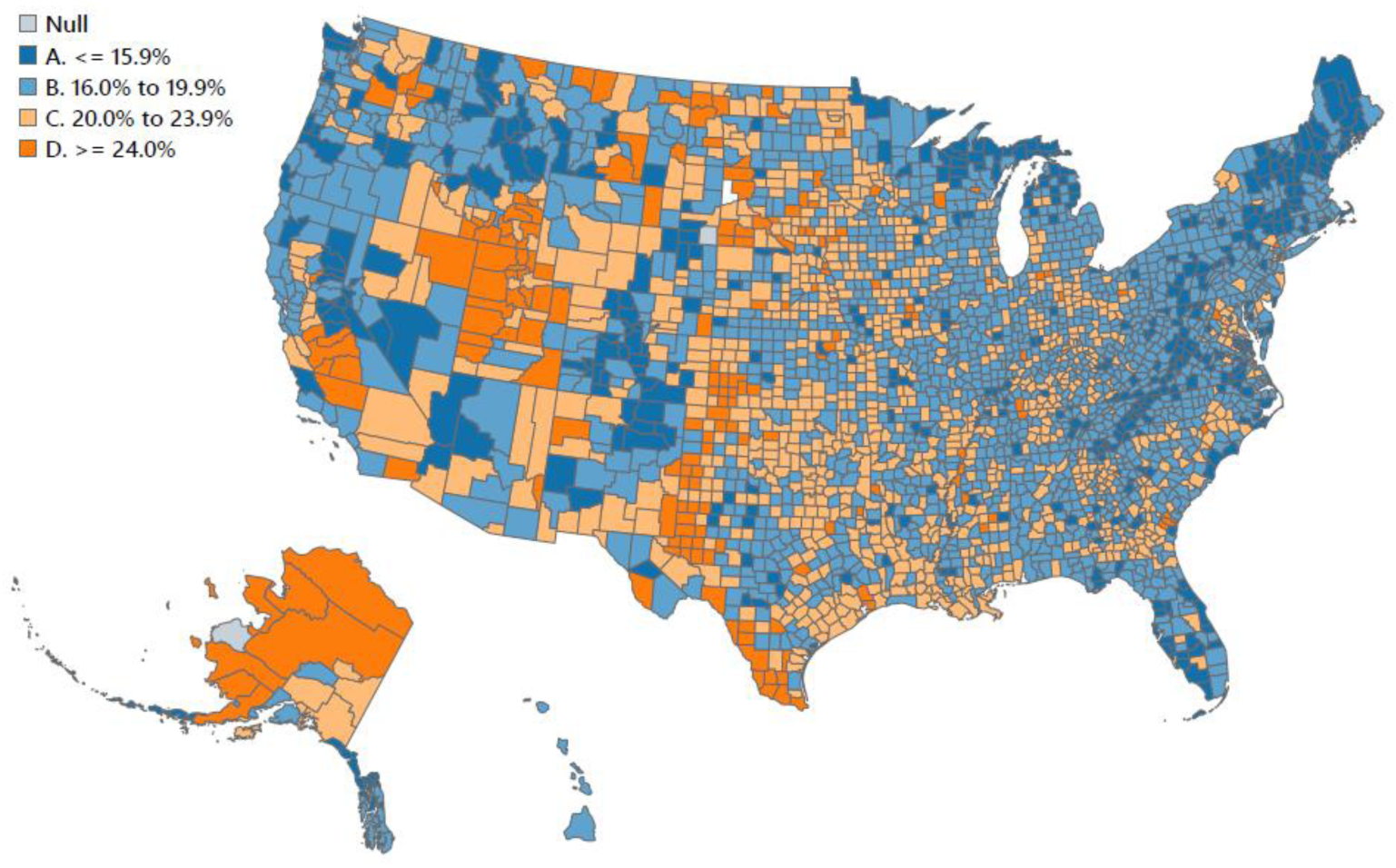
Percent of Total Population Who Are Age-Ineligible for Covid-19 Vaccine in U.S Counties. *(percent of population aged 0 to 15 years old)*

The percent of the age-eligible population who will need to be vaccinated in order to achieve 60% population vaccine coverage ranges from 71.3% in Vermont to 80.8% in Utah (Figure 3). Seventeen states, comprising 25% of the total US population, will need to vaccinate more than 75% of the age-eligible population in order to achieve 60% vaccine coverage (Figure 3). To achieve the same coverage at the county level, required vaccination rates range from 60% to 97.2%, with a median value of 74.2% and 1^st^ to 99^th^ percentile range from 68.1% to 84.5%. Local area variation is shown in Figure 4 and follows the spatial patterns observed in Figure 2 (as the rank-ordering of counties is identical for both measures).

**Figure 3.**
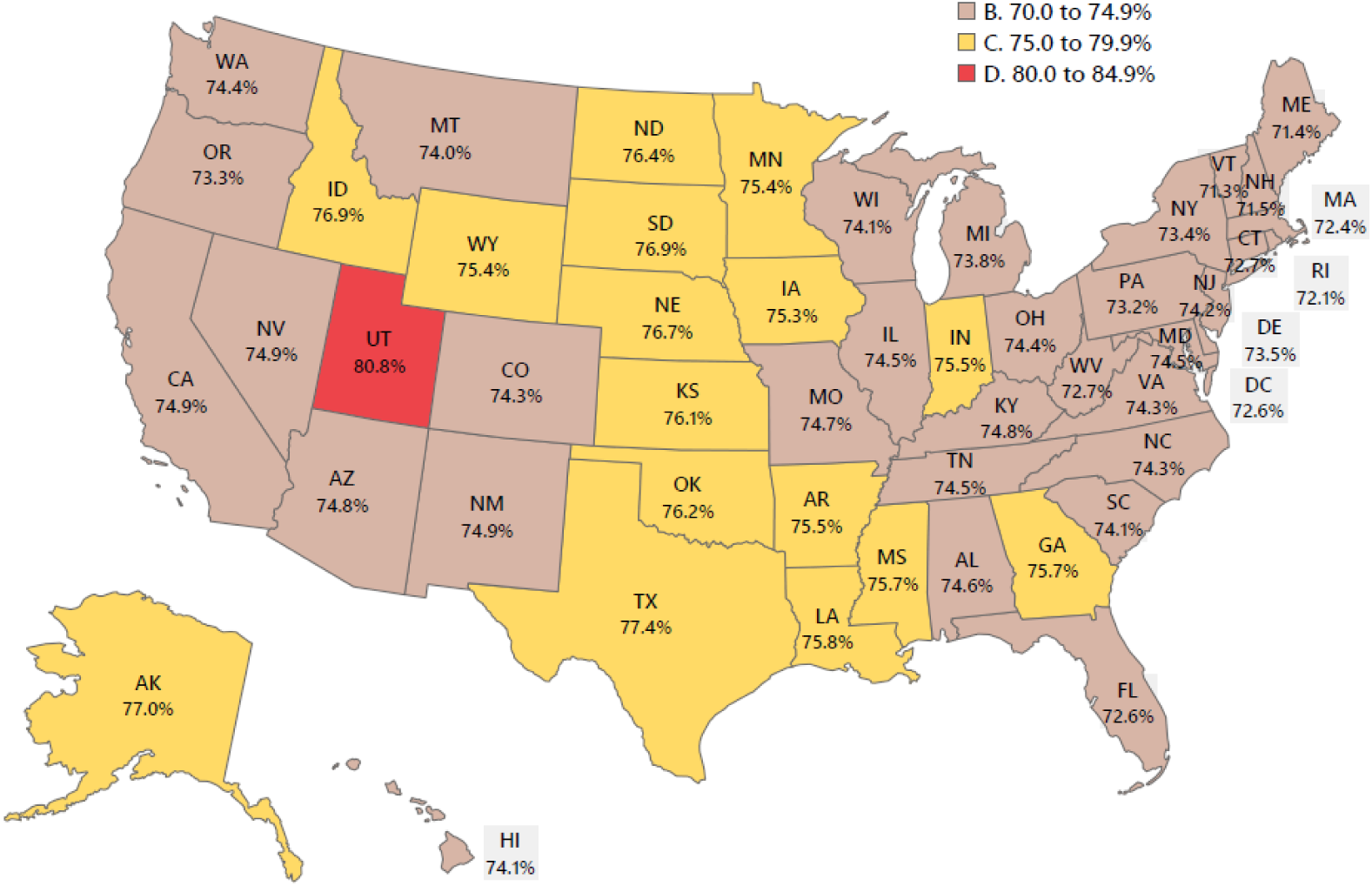
Percent if Age-Eligible Population Who Will Need to be Vaccinated to Achieve Total Population VA Vaccine Coverage of 60% in Each State.

**Figure 4.**
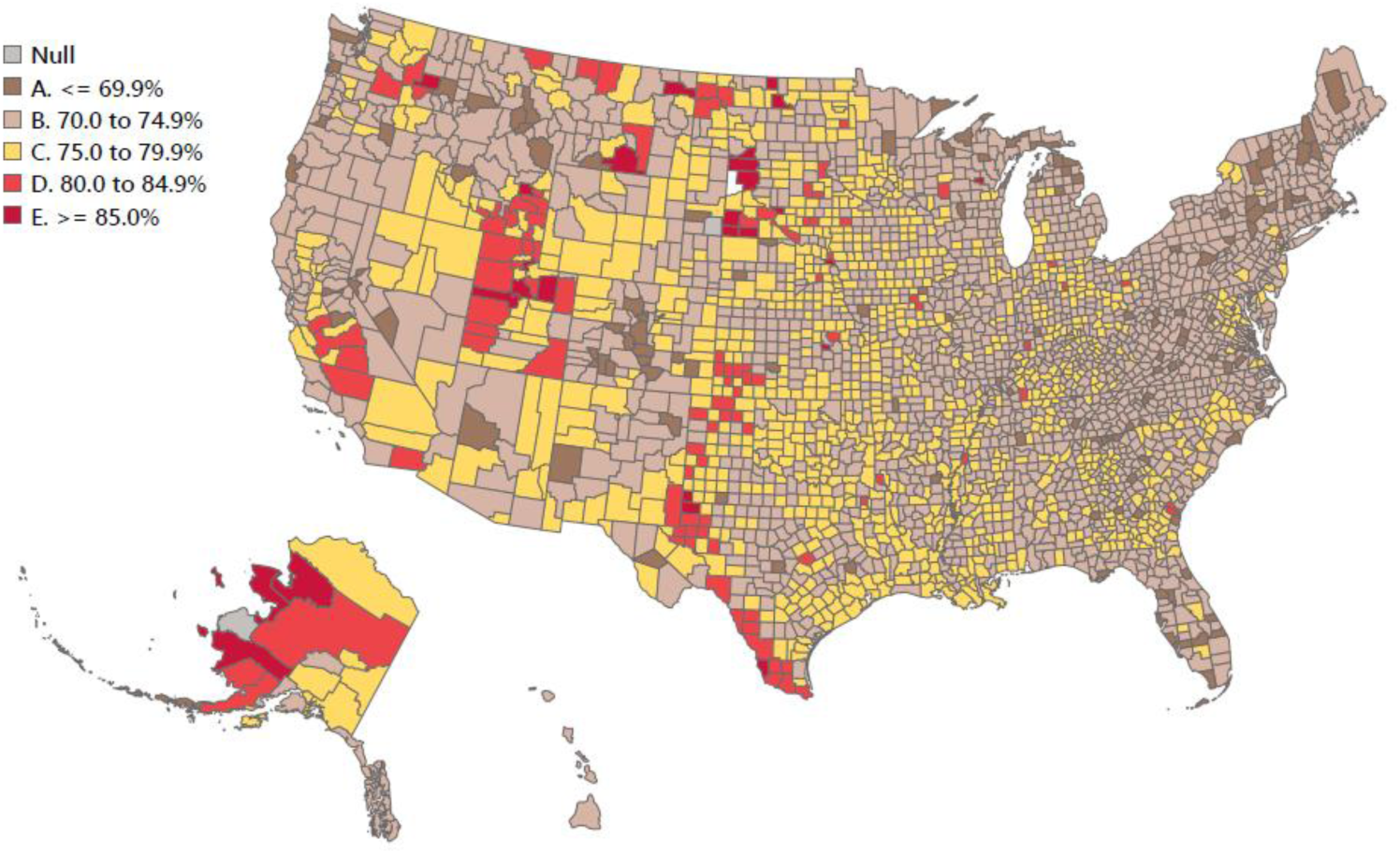
Percent of Age-Eligible Population Who Will Need to be Vaccinated to Achieve Total Population Vaccine Coverage of 60% in U.S. Counties.

Locality data are presented for the 30 largest population counties, ordered by geographic clusters (Table 1). In California, percent ineligible population in the 8 largest counties ranges from 18.2% in Oakland (Alameda County) to 23.1% in San Bernardino County. In Texas, Houston (Harris County) has the highest percent ineligible (23.6%) of any major city in the US and would need to vaccinate 78.5% of its age-eligible population in order to achieve 60% total population vaccine coverage. Results for the New York City metro counties are notable for the outlier status of Manhattan (New York County), which has one of the oldest populations of any county in the U.S., with only 13.3% of its population less than 16 years of age. In contrast, the Bronx, with 22.0% vaccine ineligible, will need to vaccinate 76.9% of its total population to achieve 60% vaccine coverage. Achievement of total population vaccine coverage of at least 75% will require vaccinating more than 90% of the population aged 16 years and older in 29 out of 30 of the largest counties in the U.S. (Table 1).

**Table 1.** Population Age-Ineligible for SARS-CoV-2 Vaccine in the 30 Largest-Population Counties, Grouped Geographically.

| County Name *(Largest City)* | National Rank in Total Population | Vaccine Ineligible Population* *(percent)* | Percent of Age-Eligible Population Needed to Vaccinate in order to Achieve Specified Target |  | Total Population *(number)* | Population 0 to 15 years old *(number)* |
| --- | --- | --- | --- | --- | --- | --- |
|  |  |  | 60% Total Coverage | 75% Total Coverage |  |  |
| California |  |  |  |  |  |  |
| Sacramento County *(Sacramento)* | 24 | 20.7 | 75.7 | 94.6 | 1,552,058 | 322,010 |
| Alameda County *(Oakland)* | 20 | 18.2 | 73.3 | 91.7 | 1,671,329 | 303,456 |
| Santa Clara County *(San Jose)* | 18 | 19.0 | 74.1 | 92.6 | 1,927,852 | 367,216 |
| Los Angeles County *(Los Angeles)* | 1 | 18.9 | 74.0 | 92.5 | 10,039,107 | 1,894,672 |
| San Bernardino County *(San Bernardino)* | 14 | 23.1 | 78.0 | 97.5 | 2,180,085 | 503,066 |
| Orange County *(Anaheim)* | 6 | 19.1 | 74.2 | 92.7 | 3,175,692 | 605,073 |
| Riverside County *(Riverside)* | 10 | 21.7 | 76.6 | 95.8 | 2,470,546 | 536,028 |
| San Diego County *(San Diego)* | 5 | 19.2 | 74.3 | 92.8 | 3,338,330 | 640,096 |
| West and Southwest |  |  |  |  |  |  |
| King County, WA *(Seattle)* | 13 | 18.0 | 73.2 | 91.5 | 2,252,782 | 405,297 |
| Clark County, NV *(Las Vegas)* | 11 | 20.4 | 75.4 | 94.2 | 2,266,715 | 461,537 |
| Maricopa County, AZ *(Phoenix)* | 4 | 20.6 | 75.6 | 94.5 | 4,485,414 | 924,936 |
| Texas |  |  |  |  |  |  |
| Tarrant County *(Ft. Worth)* | 15 | 22.9 | 77.8 | 97.3 | 2,102,515 | 480,697 |
| Dallas County *(Dallas)* | 8 | 23.1 | 78.0 | 97.5 | 2,635,516 | 609,001 |
| Harris County *(Houston)* | 3 | 23.6 | 78.5 | 98.2 | 4,713,325 | 1,110,218 |
| Bexar County *(San Antonio)* | 16 | 22.4 | 77.3 | 96.6 | 2,003,554 | 449,200 |
| Midwest |  |  |  |  |  |  |
| Wayne County, MI *(Detroit)* | 19 | 20.9 | 75.9 | 94.8 | 1,749,343 | 365,127 |
| Cook County, IL *(Chicago)* | 2 | 19.2 | 74.3 | 92.8 | 5,150,233 | 989,136 |
| Northeast |  |  |  |  |  |  |
| Middlesex County, MA *(Lowell)* | 22 | 17.2 | 72.5 | 90.6 | 1,611,699 | 277,007 |
| Philadelphia County, PA *(Philadelphia)* | 23 | 19.7 | 74.7 | 93.4 | 1,584,064 | 311,809 |
| New York City Metro |  |  |  |  |  |  |
| Bronx County *(Bronx)* | 28 | 22.0 | 76.9 | 96.2 | 1,418,207 | 312,617 |
| New York County *(Manhattan)* | 21 | 13.3 | 69.2 | 86.5 | 1,628,706 | 215,991 |
| Queens County (*Queens*) | 12 | 18.1 | 73.3 | 91.6 | 2,253,858 | 408,317 |
| Kings County (*Brooklyn*) | 9 | 20.8 | 75.8 | 94.7 | 2,559,903 | 533,082 |
| Nassau County (*Hempstead*) | 30 | 18.6 | 73.7 | 92.1 | 1,356,924 | 252,195 |
| Suffolk County (*Brookhaven*) | 26 | 18.0 | 73.2 | 91.5 | 1,476,601 | 265,634 |
| **Florida** |  |  |  |  |  |  |
| Hillsborough County (*Tampa*) | 27 | 19.6 | 74.6 | 93.3 | 1,471,968 | 288,908 |
| Orange County (*Orlando*) | 29 | 19.5 | 74.5 | 93.2 | 1,393,452 | 271,889 |
| Palm Beach County (*West Palm Beach*) | 25 | 16.7 | 72.0 | 90.0 | 1,496,770 | 249,398 |
| Broward County (*Ft. Lauderdale*) | 17 | 18.6 | 73.7 | 92.1 | 1,952,778 | 363,348 |
| Miami-Dade County (*Miami*) | 7 | 18.0 | 73.2 | 91.5 | 2,716,940 | 489,527 |

\* Vaccine ineligible population is the population aged 0-15 years old. 2019 is the latest year for which population estimates are currently available.

Vaccine eligibility varies markedly by race and ethnicity (Table 2). Non-Hispanic whites, who are 60.1% of the total U.S. population, are the oldest group, with only 16.4% vaccine-ineligible. State rates for whites range from a low of 12.9% vaccine ineligible in New Mexico, to a high of 24.3% vaccine ineligible in Utah (Table S1). There are 10 states in which fewer than 15% of the white population are vaccine ineligible. Nationally, it will require 71.8% vaccine coverage among white adults to achieve 60% total population coverage, and 89.7% vaccine coverage to achieve 75% total population coverage.

**Table 2.** Population Age-Ineligible for SARS-CoV-2 Vaccine by Race and Hispanic Ethnicity in the United States.

| Race/Ethnicity | Total<br>Population in<br>2019 | Population<br>Aged 0-15<br>years in 2019 | Percent<br>Ineligible for<br>Vaccine | Percent<br>Eligible for<br>Vaccine | Percent<br>Needed to<br>Vaccinate to<br>Achieve 60%<br>Vaccine<br>Coverage | Percent<br>Needed to<br>Vaccinate to<br>Achieve 75%<br>Vaccine<br>Coverage |
| --- | --- | --- | --- | --- | --- | --- |
| Whites (non-Hispanic) | 197,309,822 | 32,341,119 | 16.4 | 83.6 | 71.8 | 89.7 |
| Hispanics (includes all races) | 60,572,237 | 16,684,152 | 27.5 | 72.5 | 82.8 | 103.5 |
| Blacks (non-Hispanic) | 41,147,488 | 8,877,697 | 21.6 | 78.4 | 76.5 | 95.6 |
| Asians (non-Hispanic) | 18,905,879 | 3,252,220 | 17.2 | 82.8 | 72.5 | 90.6 |
| More than one race (non-Hispanic) | 7,273,281 | 2,914,001 | 40.1 | 59.9 | 100.1 | 125.1 |
| American Indians and Alaska Natives (non-Hispanic) | 2,434,908 | 546,482 | 22.4 | 77.6 | 77.4 | 96.7 |
| American Indians and Alaska Natives (Hispanic)* | 1,753,184 | 491,466 | 28.0 | 72.0 | 83.4 | 104.2 |
| Native Hawaiians and other Pacific Islanders (non-Hispanic) | 595,908 | 130,634 | 21.9 | 78.1 | 76.8 | 96.1 |
| **Everyone** | **328,239,523** | **64,746,305** | **19.7** | **80.3** | **74.7** | **93.4** |

\* NOTE: Hispanic American Indians and Alaskan Natives also appear in the totals for Hispanics listed above.

In contrast, more than one-quarter (27.5%) of Hispanics, who comprise 18.5% of the total U.S. population, are vaccine-ineligible (Table 2). Over four-fifths (82.8%) of eligible Hispanics will need to be vaccinated to achieve 60% vaccine coverage, and 75% population coverage will not be possible without child vaccinations. At the state level, there are 12 states in which more than one-third of Hispanics are age-ineligible for the COVID-19 vaccine (Table S1). More than one-quarter of Hispanics are vaccine ineligible in the two largest states: California (26.4%), and Texas (28.3%). Florida has the oldest Hispanic population, with 21.3% vaccine ineligible (Table S1).

Percent vaccine ineligible was greater than one-fifth among non-Hispanic Blacks (21.6%) (Table 2), who are 12.5% of the total US population, and ranged at the state level from a low of 17.9% in California to a high of 31.1% in Minnesota (Table S1). More than three-quarters of age-eligible Blacks will need to be vaccinated to achieve 60% population coverage, and 75% total Black population coverage will require a vaccination rate of 95.6% (Table 2).

National percent vaccine ineligible results are shown for Asians (17.2%), Non-Hispanic American Indians and Alaska Natives (22.4%), Hispanic American Indians and Alaska Natives (28.0%), and non-Hispanic Native Hawaiians and other Pacific Islanders (21.9%) in Table 2, with state results in Table S1. Persons who identify with more than one race (2.2% of the total US population) are a very young population (40.1% less than 16 years old), as a result of both secular trends in interracial childbearing, and recent changes in census data collection and reporting of racial categories.^16^

## Discussion

Historically, uptake of recommended vaccines in the general adult U.S. population has been far lower than the levels being targeted for COVID-19. Ten-year trend analyses from the CDC reveal that vaccine coverage rates for influenza, HPV, and Herpes zoster have not yet reached 50%,^17^ although these rates are slowly improving. Adult influenza vaccine coverage increased from 40.4% in the 2009-10 flu season to 48.4%, a 10-year high, in the 2019-2020 flu season.^17,18^ HPV vaccination coverage increased among women from 25.7% in 2013 to 35.3% in 2018, and among men from 7.7% to 27.7% for the same years.^19^ For the period 2010-2018, the shingles vaccine uptake remained at less than 50%, even with an amended age limit from 60 to 50 years and older in 2017.^17^ Vaccine uptake has been historically higher for the Pneumococcal vaccine, but only in the population 65 years and older. Pneumococcal vaccine coverage for elders increased from 59.7% in 2010 to 71.8% in 2018,^17^ but remained below 50% for the high-risk population aged less than 65 years.

Public health leaders are moving forward under the optimistic assumption that demand and uptake of the SARS-CoV-2 vaccines will far exceed past experience with other adult vaccines. Yet media reports on vaccine demand are fundamentally uninterpretable – lengthy queues at vaccination sites, busy phone lines, and crashing websites convey only that demand exceeds limited current capacities -- they do not provide quantitative information on the proportion of currently eligible adults who are actively attempting to access the vaccine.

Results of a survey of a representative sample of US adults conducted in December 2020 are consistent with historically low vaccination trends.^20^ Overall, only 56.2% of adults surveyed said that they would be somewhat or very likely to get a COVID-19 vaccine. Sociodemographic groups least likely to say they might get vaccinated were women (50.6%), younger adults 18-49 years (50.9%), Blacks (37.6%), Hispanics (52.7%), high school graduates with no college (47.6%), and those with some college but no 4-year degree (50.2%). Highest willingness to consider vaccination was found for the elderly >=65 years old (69.1%), Asians (80.6%), and those with a Bachelor’s degree or higher (70.3%). Pew Research Center found similar results in November 2020, with overall 60% of adults willing to get the vaccine, but only 54% of women and 42% of Blacks.^21^ The Pew study also found that among adults who typically get a flu shot, 77% were willing to get the COVID-19 vaccine, contrasted with only 38% of those who rarely have taken flu shots in the past.

A recent expert commentary in the Lancet explained many of the challenges to achieving herd immunity to SARS-CoV-2 through vaccination.^22^ However, the impact of the vaccine-ineligible (child) population was not mentioned or acknowledged. It is well understood that herd immunity parameters are always disease-specific and population-contingent.^23-25^ Herd immunity depends on both vaccine-conferred immunity and naturally-acquired immunity among persons who have recovered from SARS-CoV-2 infections.^23^ The emergence in late 2020/early 2021 of new SARS-CoV-2 variants that are proving to be more transmissible^26^ has resulted in experts revising upward the percent of the total population who will need to achieve individual immunity (through either vaccination or illness recovery) before herd immunity will protect those who are not immune. ^23^

Over 3.2 million children and teens aged 0-19 years have ever tested positive for SARS-CoV-2 in the U.S.^8^ An optimistic assumption is that many of these children have achieved long-lasting natural immunity. However, due to critical deficits in surveillance of COVID-19 in children, there is little evidence with which to test this assumption. Two fundamental questions germane to the goal of achieving herd immunity remain unanswered: (1) how long-lasting is vaccine-conferred individual immunity? and (2) how long-lasting is infection-conferred individual immunity? Preliminary research on cellular immunity from natural infection suggests the possibility of lasting protection;^27,28^ however, immune memory from infection has proven variable. In the absence of robust longitudinal data, a clear understanding of the mechanisms of humoral and cellular immune responses remains incomplete^29^ Because of these uncertainties, the CDC currently recommends that everyone who is age-eligible should be vaccinated, including individuals who have previously tested positive for the virus and/or survived clinical COVID-19 illness.^30^

Differences in the age structure of racial/ethnic population groups will impact the success of COVID-19 vaccination programs. Hispanics, Blacks, American Indians, and Alaska Natives have the highest percent of population who are age-ineligible for vaccine. These are the same groups who have suffered the highest burden of COVID-19 incidence^31-36^ and mortality.^37-40^ In national surveys, Blacks and Hispanics reported greater vaccine hesitancy than Asians or non-Hispanic whites.^20,21^ Moreover, early data on vaccine rollout reveal pervasive barriers and lower vaccination rates in Black and Hispanic communities.^41-43^

For these reasons, vaccination campaigns will almost certainly *increase* racial/ethnic disparities in the incidence of COVID-19. The groups with the lowest COVID-19 incidence rates in the pre-vaccine era (non-Hispanic whites and Asians) also have the smallest percent vaccine ineligible populations and will benefit the most from adult vaccination programs. Conversely, the persistence of racial/ethnic segregation in housing, occupations, religious communities, and other social institutions in 21^st^ century America,^44^ coupled with higher levels of vaccine ineligibility and vaccine hesitancy, creates the conditions for endemic entrenchment of COVID-19 in many Hispanic, Black, and Native communities.

## CONCLUSIONS

The rapid development, testing, and mass production of essential COVID-19 vaccines has been groundbreaking, and their proven short-term efficacy and safety have inspired hope in a pandemic-weary world. Yet our analyses show that the vaccine-ineligibility of most children for at least the next 1-2 years means that achievement of high levels of vaccine coverage will be very unlikely in many vulnerable geographic areas and for Hispanics, Blacks, American Indians, and other racial/ethnic minority groups. Moreover, the recent emergence of new SARs-CoV-2 strains with greater transmissibility^26^ and the potential to partially dodge vaccine-conferred immunity^45^ will further impede public health efforts to control the pandemic.

There remain immense challenges to adult vaccine uptake that must be addressed as efforts move forward to reach, persuade, and vaccinate a high proportion of the US population. The historically low rates of adult coverage for routine, age-appropriate vaccines underscore the need for culturally sensitive health education efforts to overcome widespread fear and vaccine hesitancy. At the same time, clinical trials for efficacy and safety of child vaccines must become a global priority. Finally, overcoming vaccine hesitancy among women, particularly mothers, must be recognized as an urgent and vital necessity.^46^ This is because women are typically the primary family healthcare consumers,^47,48^ and once pediatric COVID-19 vaccines become available, mothers will control child uptake to a large extent.^49,50^

## Data Availability

All data analyzed in this study are in the public domain and available from CDC WONDER and the US Census Bureau.

