## Supplemental Table for "Population Age-Ineligible for COVID-19 Vaccine in the United States: Implications for State, County, and Race/Ethnicity Vaccination Targets"

| Table S1. Percent Vaccine Ineligible by Race/Ethnicity and State, Based on 2019 Population Estimates | | | | | | | | | | | | | | |
|---|---|---|---|---|---|---|---|---|---|---|---|---|---|---|
| | Percent Vaccine Ineligible (Ages 0 to 15 Years Old) | | | | | | | All Ages Population | | | | | | |
| State | White | Hispanic | Black | Asian | American Indian/ Alaska Native | Native Hawaiian/ other Pacific Islander | More Than One Race | White | Hispanic | Black | Asian | American Indian/ Alaska Native | Native Hawaiian/ other Pacific Islander | More Than One Race |
| Alabama | 17.3 | 36.4 | 21.4 | 19.1 | 18.6 | 21.8 | 43.1 | 3,200,828 | 223,278 | 1,297,775 | 72,046 | 34,799 | 2539 | 78,990 |
| Alaska | 17.9 | 30.7 | 20.3 | 18.3 | 29.2 | 28.2 | 40.3 | 440,078 | 53,212 | 24,190 | 46,606 | 113,953 | 10012 | 49,564 |
| Arizona | 14.2 | 28.1 | 22.2 | 16.5 | 27.5 | 19.4 | 38.6 | 3,939,690 | 2,310,590 | 325,777 | 249,264 | 385,903 | 13913 | 152,502 |
| Arkansas | 17.9 | 33.6 | 23.6 | 20.2 | 22.3 | 32.4 | 42.5 | 2,173,848 | 236,631 | 465,209 | 48,588 | 30,717 | 10617 | 59,740 |
| California | 14.0 | 26.4 | 17.9 | 15.5 | 21.2 | 19.5 | 35.5 | 14,423,748 | 15,574,880 | 2,221,363 | 5,869,038 | 649,862 | 146449 | 1,114,409 |
| Colorado | 16.0 | 28.0 | 21.0 | 17.8 | 24.1 | 21.0 | 37.2 | 3,896,103 | 1,256,904 | 233,523 | 192,733 | 92,982 | 7975 | 134,185 |
| Connecticut | 14.4 | 27.3 | 19.9 | 18.9 | 23.8 | 22.6 | 39.9 | 2,350,123 | 600,955 | 368,834 | 172,355 | 20,567 | 1266 | 63,948 |
| Delaware | 14.5 | 32.3 | 21.3 | 18.6 | 21.8 | 23.1 | 43.0 | 600,349 | 93,391 | 214,658 | 39,174 | 6,564 | 351 | 22,838 |
| District of Columbia | 10.8 | 25.5 | 19.5 | 9.3 | 22.8 | 17.1 | 30.6 | 264,400 | 79,477 | 313,290 | 30,541 | 4,130 | 339 | 16,289 |
| Florida | 13.5 | 21.3 | 22.4 | 15.8 | 18.4 | 17.8 | 40.0 | 11,436,685 | 5,663,860 | 3,335,263 | 610,090 | 108,950 | 13827 | 363,997 |
| Georgia | 17.3 | 32.1 | 22.1 | 19.3 | 24.8 | 23.2 | 43.6 | 5,523,358 | 1,048,724 | 3,359,449 | 454,147 | 56,008 | 6641 | 200,720 |
| Hawaii | 13.0 | 33.8 | 18.6 | 11.9 | 15.7 | 21.3 | 29.8 | 306,622 | 150,864 | 27,780 | 515,909 | 5,570 | 133369 | 278,311 |
| Idaho | 20.3 | 32.3 | 27.3 | 17.1 | 28.1 | 22.3 | 38.5 | 1,458,277 | 229,490 | 13,584 | 25,778 | 31,120 | 3174 | 37,024 |
| Illinois | 16.5 | 27.9 | 21.1 | 17.6 | 19.9 | 21.1 | 42.3 | 7,702,651 | 2,219,882 | 1,783,443 | 731,524 | 75,500 | 3717 | 211,285 |
| Indiana | 18.4 | 32.7 | 24.3 | 20.2 | 21.6 | 24.7 | 46.0 | 5,278,982 | 489,353 | 645,375 | 171,203 | 28,340 | 2577 | 129,041 |
| Iowa | 18.4 | 34.3 | 29.1 | 20.2 | 31.5 | 29.2 | 48.0 | 2,682,696 | 198,550 | 122,388 | 82,604 | 17,060 | 4115 | 55,044 |
| Kansas | 18.7 | 32.7 | 23.1 | 18.9 | 23.3 | 25.6 | 42.7 | 2,196,863 | 356,073 | 167,325 | 90,552 | 35,063 | 2818 | 76,412 |
| Kentucky | 18.4 | 34.1 | 22.3 | 22.0 | 20.2 | 24.2 | 46.4 | 3,759,463 | 174,706 | 367,899 | 69,783 | 13,404 | 3074 | 83,352 |
| Louisiana | 18.1 | 29.8 | 23.3 | 18.5 | 22.4 | 19.7 | 45.8 | 2,715,282 | 246,972 | 1,500,991 | 81,660 | 36,596 | 1840 | 71,798 |
| Maine | 15.4 | 27.3 | 30.6 | 15.6 | 22.1 | 24.8 | 37.8 | 1,249,597 | 23,700 | 21,554 | 17,083 | 9,767 | 375 | 22,913 |
| Maryland | 16.0 | 31.1 | 20.0 | 18.0 | 25.6 | 19.4 | 42.1 | 3,025,781 | 643,822 | 1,810,267 | 398,519 | 36,840 | 3025 | 149,504 |
| Massachusetts | 14.5 | 27.2 | 21.1 | 17.3 | 20.4 | 20.1 | 38.9 | 4,897,800 | 854,907 | 505,757 | 490,023 | 34,310 | 2989 | 128,619 |
| Michigan | 16.8 | 30.7 | 22.2 | 18.7 | 22.1 | 19.4 | 40.8 | 7,464,662 | 528,205 | 1,374,886 | 332,320 | 73,865 | 2814 | 226,706 |
| Minnesota | 17.6 | 33.5 | 31.1 | 24.3 | 32.2 | 27.1 | 45.6 | 4,460,149 | 315,130 | 382,621 | 288,583 | 77,479 | 2941 | 129,957 |
| Mississippi | 18.1 | 31.8 | 23.1 | 18.2 | 27.4 | 19.4 | 46.1 | 1,678,232 | 100,110 | 1,113,643 | 32,251 | 18,705 | 1042 | 35,804 |
| Missouri | 18.1 | 32.0 | 23.0 | 18.6 | 22.2 | 27.7 | 42.9 | 4,857,512 | 268,708 | 711,702 | 130,717 | 35,839 | 8246 | 133,445 |
| Montana | 17.1 | 31.9 | 22.7 | 14.7 | 32.3 | 16.9 | 35.9 | 917,711 | 43,289 | 5,766 | 9,412 | 71,096 | 767 | 26,880 |
| Nebraska | 19.0 | 35.3 | 26.8 | 22.3 | 37.4 | 24.0 | 46.3 | 1,513,172 | 219,645 | 94,830 | 51,496 | 29,285 | 1229 | 37,703 |
| Nevada | 14.3 | 28.2 | 22.7 | 13.1 | 23.6 | 20.5 | 39.1 | 1,483,933 | 900,600 | 286,466 | 254,474 | 51,946 | 19718 | 108,602 |
| New Hampshire | 15.4 | 27.8 | 22.4 | 18.5 | 17.7 | 19.1 | 37.2 | 1,220,437 | 54,589 | 20,054 | 39,797 | 4,114 | 403 | 21,507 |
| New Jersey | 16.0 | 26.0 | 19.9 | 18.9 | 21.2 | 23.0 | 40.3 | 4,850,995 | 1,856,844 | 1,148,564 | 870,472 | 55,429 | 3163 | 139,329 |
| New Mexico | 12.9 | 24.6 | 18.8 | 14.6 | 26.3 | 16.3 | 34.2 | 772,630 | 1,032,942 | 39,583 | 32,698 | 229,794 | 1371 | 32,372 |
| New York | 15.8 | 24.1 | 18.9 | 16.7 | 20.0 | 19.8 | 39.0 | 10,755,420 | 3,751,058 | 2,813,773 | 1,713,428 | 189,748 | 8983 | 354,118 |
| North Carolina | 15.9 | 34.1 | 20.5 | 20.8 | 25.4 | 23.5 | 43.7 | 6,567,102 | 1,025,830 | 2,240,609 | 322,955 | 165,597 | 6967 | 207,402 |
| North Dakota | 19.1 | 35.7 | 28.9 | 19.9 | 34.2 | 25.6 | 44.9 | 637,513 | 31,532 | 24,866 | 12,561 | 42,465 | 524 | 15,855 |
| Ohio | 17.5 | 31.9 | 23.5 | 19.7 | 21.3 | 25.1 | 44.5 | 9,169,092 | 470,462 | 1,481,352 | 287,255 | 34,010 | 4824 | 253,171 |
| Oklahoma | 17.3 | 34.6 | 22.6 | 18.9 | 27.0 | 27.4 | 36.9 | 2,572,388 | 438,110 | 293,458 | 91,286 | 371,104 | 7091 | 223,903 |
| Oregon | 15.4 | 30.4 | 22.3 | 15.6 | 24.4 | 20.8 | 34.4 | 3,165,484 | 566,847 | 82,610 | 198,697 | 77,115 | 17074 | 139,868 |

| Table S1. Percent Vaccine Ineligible by Race/Ethnicity and State, Based on 2019 Population Estimates | | | | | | | | | | | | | | |
|---|---|---|---|---|---|---|---|---|---|---|---|---|---|---|
| | Percent Vaccine Ineligible (Ages 0 to 15 Years Old) | | | | | | | All Ages Population | | | | | | |
| **State** | **White** | **Hispanic** | **Black** | **Asian** | **American Indian/ Alaska Native** | **Native Hawaiian/ other Pacific Islander** | **More Than One Race** | **White** | **Hispanic** | **Black** | **Asian** | **American Indian/ Alaska Native** | **Native Hawaiian/ other Pacific Islander** | **More Than One Race** |
| Pennsylvania | 15.8 | 30.2 | 21.9 | 19.0 | 26.4 | 21.6 | 43.9 | 9,693,578 | 1,000,150 | 1,394,630 | 471,559 | 50,687 | 3936 | 219,290 |
| Rhode Island | 13.4 | 28.0 | 21.1 | 17.3 | 31.5 | 21.5 | 38.4 | 755,931 | 172,644 | 64,966 | 37,922 | 11,453 | 659 | 22,639 |
| South Carolina | 16.4 | 32.3 | 21.2 | 18.4 | 22.8 | 22.3 | 45.7 | 3,277,542 | 307,118 | 1,360,342 | 91,375 | 28,236 | 3101 | 90,248 |
| South Dakota | 19.1 | 38.2 | 30.3 | 21.8 | 36.1 | 29.0 | 46.6 | 721,053 | 37,351 | 19,447 | 13,393 | 80,004 | 525 | 19,413 |
| Tennessee | 17.2 | 35.7 | 22.2 | 19.3 | 24.1 | 22.6 | 44.1 | 5,019,540 | 391,382 | 1,141,790 | 130,143 | 32,661 | 3762 | 122,860 |
| Texas | 17.2 | 28.3 | 22.2 | 19.5 | 21.6 | 23.1 | 41.6 | 11,950,774 | 11,525,578 | 3,501,610 | 1,457,549 | 294,902 | 25861 | 440,341 |
| Utah | 24.3 | 32.5 | 27.6 | 17.7 | 30.7 | 27.6 | 44.2 | 2,493,759 | 462,051 | 38,056 | 81,646 | 49,720 | 31393 | 68,652 |
| Vermont | 15.4 | 22.9 | 22.9 | 17.2 | 15.0 | 18.3 | 34.5 | 577,539 | 12,719 | 8,152 | 11,813 | 2,441 | 197 | 11,511 |
| Virginia | 16.7 | 29.1 | 20.0 | 18.3 | 19.6 | 19.6 | 42.2 | 5,227,904 | 834,422 | 1,632,226 | 578,504 | 46,602 | 6541 | 233,128 |
| Washington | 16.0 | 32.5 | 21.9 | 16.3 | 28.8 | 23.2 | 38.1 | 5,140,589 | 991,721 | 304,224 | 711,626 | 146,799 | 54633 | 315,594 |
| West Virginia | 17.0 | 29.2 | 18.3 | 15.4 | 15.9 | 18.5 | 43.6 | 1,648,512 | 31,162 | 62,775 | 14,393 | 4,585 | 411 | 31,058 |
| Wisconsin | 16.6 | 33.1 | 26.8 | 23.7 | 28.7 | 24.7 | 45.6 | 4,709,065 | 413,208 | 372,273 | 172,205 | 68,628 | 2318 | 100,929 |
| Wyoming | 18.8 | 30.8 | 21.6 | 14.9 | 30.9 | 18.7 | 39.5 | 484,380 | 58,609 | 6,520 | 6,129 | 15,778 | 412 | 10,511 |
